# SARS-CoV-2 infection induces autoimmune antibody secretion more in lean than in obese COVID-19 patients

**DOI:** 10.1101/2021.05.05.21256686

**Authors:** Daniela Frasca, Lisa Reidy, Maria Romero, Alain Diaz, Carolyn Cray, Kristin Kahl, Bonnie B. Blomberg

## Abstract

**Background/Objectives:** Obesity decreases the secretion of SARS-CoV-2-specific IgG antibodies in the blood of COVID-19 patients. How obesity impacts the secretion of autoimmune antibodies in COVID-19 patients, however, is not understood. The serum of adult COVID-19 patients contains autoimmune antibodies generated in response to virus-induced tissue damage and cell death leading to the release of intracellular antigens not known to be immunogenic autoantigens. The objective of this study is to evaluate the presence of autoimmune antibodies in COVID-19 patients with obesity.

**Subjects/Methods:** Thirty serum samples from individuals who tested positive for SARS-CoV-2 infection by RT-PCR were collected from inpatient and outpatient settings. Of these, 15 were lean (BMI<25), and 15 were obese (BMI ≥30). Control serum samples were from 30 uninfected individuals, age-gender- and BMI-matched, recruited before the current pandemic. Serum IgG antibodies against two autoimmune specificities, as well as against SARS-CoV-2 Spike protein, were measured by ELISA. IgG autoimmune antibodies were specific for malondialdehyde (MDA), a marker of oxidative stress and lipid peroxidation, and for adipocyte-derived protein antigens (AD), markers of virus-induced cell death in the obese AT.

**Results:** Our results show that SARS-CoV-2 infection induces anti-MDA and anti-AD autoimmune antibodies more in lean than in obese patients as compared to uninfected controls. Serum levels of these autoimmune antibodies, however, are always higher in obese versus lean COVID-19 patients. Moreover, because the autoimmune antibodies found in serum samples of COVID-19 patients have been correlated with serum levels of C-reactive protein (CRP), a general marker of inflammation, we also evaluated the association of anti-MDA and anti-AT antibodies with serum CRP and found a significant association between CRP and autoimmune antibodies in our cohort of lean and obese COVID-19 patients.

**Conclusions:** Our results highlight the importance of evaluating the quality of the antibody response in COVID-19 patients with obesity, particularly the presence of autoimmune antibodies, and identify biomarkers of self-tolerance breakdown. This is crucial to protect this vulnerable population that is at higher risk of responding poorly to infection with SARS-CoV-2 compared to lean controls.

## Introduction

The novel single-stranded RNA coronavirus SARS-CoV-2 (Severe Acute Respiratory Syndrome Corona Virus-2) emerged in the last months of 2019, caused the worldwide COVID-19 pandemic, and was responsible for different clinical manifestations ranging from mild disease to severe respiratory tract infections, multiorgan failure, and death. The severe manifestations of the disease are associated with an exuberant inflammatory response and the development of a condition known as cytokine storm (1). Published data have indicated that inflammaging, the chronic low-grade systemic inflammation (2), is a major cause of the cellular and molecular changes induced by SARS-CoV-2 and can be responsible for the highest number of deaths (3). Inflammaging induces chronic immune activation (IA) and dysfunctional immunity (4).

Resolution of SARS-CoV-2 infection requires both innate and adaptive immune responses that lead to the clearance and elimination of the virus from the organism. B cells contribute to this process by producing neutralizing antibodies that prevent the spread of infectious virions, control virus dissemination, and reduce tissue damage. Neutralizing antibodies generated against the Spike glycoprotein of the SARS-CoV-1 in the 2002-2003 pandemic have shown efficacy in protecting from severe disease (5). Moreover, during the current pandemic, it has been shown that neutralizing antibodies against the Spike glycoprotein of SARS-CoV-2, found in plasma from convalescent COVID-19 patients, induced fast recovery when transfused into critically ill patients (6-8).

Obesity is an inflammatory condition associated with inflammaging and chronic IA, contributing to functional impairment of immune cells, and decreased immunity. Obese individuals have been shown to respond poorly to infections (9-11), vaccination (12-14), and therapies, e.g., for autoimmune conditions (15). Obesity and associated inflammation lead to several debilitating chronic diseases such as type-2 diabetes mellitus (16-18), cardiovascular disease (19), cancer (20), atherosclerosis (21), psoriasis (22), Alzheimer’s disease (23, 24), and inflammatory bowel disease (25). Therefore, obesity represents an additional risk factor for COVID-19 patients. A strong association between obesity, obesity-associated comorbidities, and severe outcomes of COVID-19 has indeed been shown (26), with adult COVID-19 symptomatic patients with Body Mass Index (BMI) >30 showing higher admission to acute and critical care compared to lean and overweight individuals (BMI <30) (27). It has been proposed that the obese adipose tissue (AT) in the thorax and abdominal areas is heavily infiltrated with immune cells (28, 29). These, once activated by the SARS-CoV-2 virus, would fuel local inflammation and exacerbate inflammaging through the secretion of additional pro-inflammatory mediators that can further compromise lung function (30, 31). AT may also be a viral reservoir, playing a crucial role in maintaining local and systemic inflammation, persistent IA, and immune dysfunction (32).

We have previously evaluated the effects of obesity on the secretion of SARS-CoV-2-specific IgG antibodies in the blood of lean and overweight/obese COVID-19 patients, and we have shown that SARS-CoV-2 IgG antibodies are negatively associated with BMI in COVID-19 patients, as expected based on the known effects of overweight/obesity on humoral immunity (33). In this study, we have measured the presence of antibodies with autoimmune specificities in the sera of lean (BMI <25) and obese (BMI>30) COVID-19 patients, as compared to age-, gender- and BMI-matched uninfected controls, without previous history of autoimmunity. This study included uninfected controls since obesity per se is associated with the secretion of autoimmune antibodies, as previously demonstrated (28, 34, 35). It has been recently shown that the sera of adult COVID-19 patients contain antibodies with autoimmune specificities (36). We hypothesized that SARS-CoV-2 infection in COVID-19 patients induces oxidative stress and tissue damage, leading to cell death and release of intracellular antigens that are not known to be immunogenic autoantigens, and more in obese than in lean patients. Therefore, this study evaluated a cohort of lean and obese COVID-19 patients for the presence of autoimmune antibodies specific for malondialdehyde (MDA), which is used as a marker of oxidative stress and lipid peroxidation. In addition, adipocyte-derived protein antigens (AD), established markers of cell death in the obese AT, were also evaluated. In contrast to the study’s hypothesis, the results show that SARS-CoV-2 infection induces MDA- and AD-specific autoimmune antibodies more in lean than in obese patients as compared to uninfected controls, but with serum levels of these autoimmune antibodies always higher in obese patients. Moreover, because autoimmune antibodies in serum samples of COVID-19 patients are correlated with serum levels of inflammation-associated C-reactive protein (CRP) (36), we also evaluated the association of anti-MDA and anti-AD IgG with serum CRP. CRP is a marker of pathogen-driven pulmonary inflammation, embolism, and disseminated intravascular coagulation, all characteristics of COVID-19, and therefore CRP is a predictive marker of adverse health outcomes (37-40). We found a significant association between CRP and autoimmune antibodies in our cohort of lean and obese COVID-19 patients.

## Methods

### Participants

Experiments were performed using serum samples isolated from individuals who tested positive for SARS-CoV-2 RNA by reverse transcriptase-polymerase chain reaction (RT-PCR) of nasopharyngeal swab samples. Only serum samples were collected from patients, and samples used in this study were only taken for routine clinical care purposes. Samples were de-identified before being used in this study. In total, 30 serum samples from individuals tested positive for SARS-CoV-2 infection by RT-PCR, 15 lean (BMI<25) and 15 obese (BMI ≥30), were collected from both inpatient and outpatient settings and frozen until testing was performed. Both lean and obese patients had multiple respiratory symptoms (high fever, cough, shortness of breath, hypoxia), as evaluated at the time of hospital admission, but none was admitted to ICU. The research was approved and reviewed by the Institutional Review Board (IRB, protocol #20200504, PI Dr. Reidy) at the University of Miami, which reviews all human research conducted under the auspices of the University of Miami.

Control serum samples were from 30 uninfected individuals, age-gender- and BMI-matched, recruited at the University of Miami before the current pandemic (IRB, protocols #20070481 and #20160542, PI Dr. Frasca). Both lean and obese uninfected controls were screened for diseases known to alter the immune response or for consumption of medications that could alter the immune response. We excluded subjects with autoimmune diseases, congestive heart failure, cardiovascular disease, chronic renal failure, malignancies, renal or hepatic diseases, infectious disease, trauma or surgery, pregnancy, or documented current substance and/or alcohol abuse.

The characteristics of COVID-19 patients and controls are in Table 1. Both COVID-19 patients and uninfected controls were without prior history of autoimmunity.

**Table 1.** Characteristics of enrolled participants

|  | Uninfected |  | SARS-CoV-2-infected |  |
| --- | --- | --- | --- | --- |
|  | Lean | Obese | Lean | Obese |
| Age, mean±SE | 53±4 | 50±2 | 64±9 | 55±4 |
| BMI, mean±SE | 22±1 | 37±2 | 22±3 | 35±2 |
| Males | 7 | 8 | 7 | 6 |
| Females | 8 | 7 | 8 | 9 |

### SARS-CoV-2 tests

SARS-CoV-2 RNA was detected by RT-PCR of nasopharyngeal swab samples. RT-PCR tests were performed at the clinical laboratories of the Department of Pathology & Laboratory Medicine using either the Diasorin or the BDMax assay and reagents. Depending on submission type (symptomatic or asymptomatic) the sample was assigned and tested per manufacturer guidelines.

### ELISA to measure Spike-specific IgG antibodies

Serum IgG antibodies against SARS-CoV-2 Spike protein were measured by an ELISA developed and standardized in our laboratory. Briefly, 96-well microplates (Immulon 4HBX, Thermo Scientific) were coated with recombinant NCP-CoV (2019-nCoV) Spike protein (S1+S2 ECD) (Sino Biological #40589-V08B1) at 2 µg/mL for 1 hr at room temperature. Plates were then washed with Tween-20 0.05% in PBS (PBST) and blocked with assay buffer (1% BSA in PBS) for 1 hr at 37°C. After blocking, all subsequent steps were performed by a DYNEX DS2® Automated ELISA system (DYNEX Technologies, Chantilly, VA, USA). Serum samples diluted 1:50,000 in assay buffer were added in duplicate, and plates were incubated for 2 hrs. Plates were washed with PBST and 100 µL per well of a peroxidase-conjugated goat anti-human IgG (Jackson ImmunoResearch #109-036-098), diluted 1:10,000 in assay buffer, were added. After 1 hr incubation, plates were washed, and a stabilized 3,3′,5,5′-Tetramethylbenzidine (TMB) substrate (Sigma) was added to the wells. The enzymatic reaction was stopped after 20 min with a Stop solution (1 M sulfuric acid), and absorbance at 450 nm was read by the DYNEX DS2 instrument.

### ELISA to measure autoimmune antibodies

To measure MDA-specific IgG antibodies, ELISA plates were coated with MDA-modified Bovine Serum Albumin protein (MDA-BSA, MyBioSource MBS390120), at the concentration of 10 µg/mL. Control ELISA plates were coated with BSA at a concentration of 10 µg/mL. MDA-specific OD values were calculated by subtracting BSA OD values from MDA-BSA OD values.

To measure AD-specific IgG antibodies, we isolated the adipocytes from the subcutaneous adipose tissue of patients undergoing weight reduction surgeries (bilateral breast reduction), as previously described (28). After isolation, the adipocytes were centrifuged in a 5415C Eppendorf microfuge (2,000 rpm, 5 min). Total cell lysates were obtained using the M-PER (Mammalian Protein Extraction Reagent, ThermoFisher), according to the manufacturer’s instructions. Aliquots of the protein extracts were stored at -80°C. Protein content was determined by Bradford (41). Adipocyte-derived protein lysates (AD) at a concentration of 10 µg/mL were used to coat ELISA plates.

For all tests, serum samples were diluted 1:1000 with sample buffer.

### ELISA to measure serum C-reactive protein (CRP)

Serum levels of CRP were measured using the commercially available kit R&D # DCRP00.

### Statistical analyses

To examine differences between 4 groups, one-way ANOVA was used. Group-wise differences were analyzed afterward with Bonferroni’s multiple comparisons test, with p<.05 set as a criterion for significance. To examine differences between the 2 groups, Student’s t-tests (two-tailed) were used. To examine relationships between variables, bivariate Pearson’s correlation analyses were performed. GraphPad Prism version 8.4.3 software was used to construct all graphs.

## Results

### Evaluation of Spike-specific IgG antibodies in serum samples of lean and obese COVID-19 patients, as compared to uninfected controls

The results for Spike-specific IgG antibodies in serum samples of 30 COVID-19 patients and 30 uninfected controls, age-gender- and BMI-matched, are shown in Fig. 1. Antibodies were measured by an ELISA previously developed and standardized in our laboratory (33). Results show significantly lower levels of Spike-specific IgG antibodies in obese versus lean COVID-19 patients, confirming our published findings that Spike-specific IgG antibodies in serum are negatively associated with BMI in COVID-19 patients (33). Spike-specific IgG antibodies were detected at extremely low levels in serum samples isolated from uninfected lean and obese age-gender- and BMI-matched participants recruited before the pandemic.

**Figure 1.**
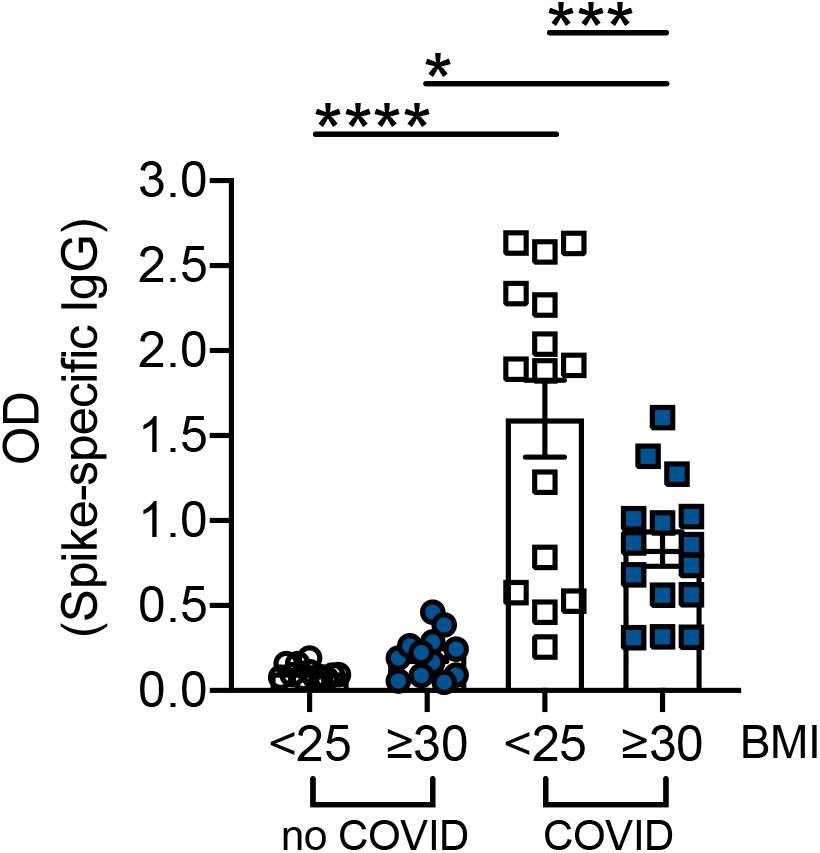
Evaluation of Spike-specific IgG antibodies in serum samples of lean and obese COVID-19 patients, as compared to uninfected controls. SARS-CoV-2 Spike-specific IgG antibodies were measured by ELISA. *p<0.05, ***p<0.001, ****p<0.0001.

### Evaluation of autoimmune IgG antibodies in serum samples of lean and obese COVID-19 patients, as compared to uninfected controls

We evaluated the presence of IgG antibodies with two autoimmune specificities in serum samples of lean and obese COVID-19 patients as compared to uninfected controls. Results in Fig. 2 show that SARS-CoV-2 infection significantly increases serum levels of anti-MDA IgG antibodies (A) and anti-AD IgG antibodies (B) in both lean and obese patients, with a greater increase in lean versus obese patients. Fold-increases in lean versus obese were 3.1 versus 1.6 for anti-MDA IgG antibodies and 6.4 versus 1.5 for anti-AD IgG antibodies. However, serum levels of all these antibodies were always higher in obese versus lean COVID-19 patients and uninfected controls.

**Figure 2.**
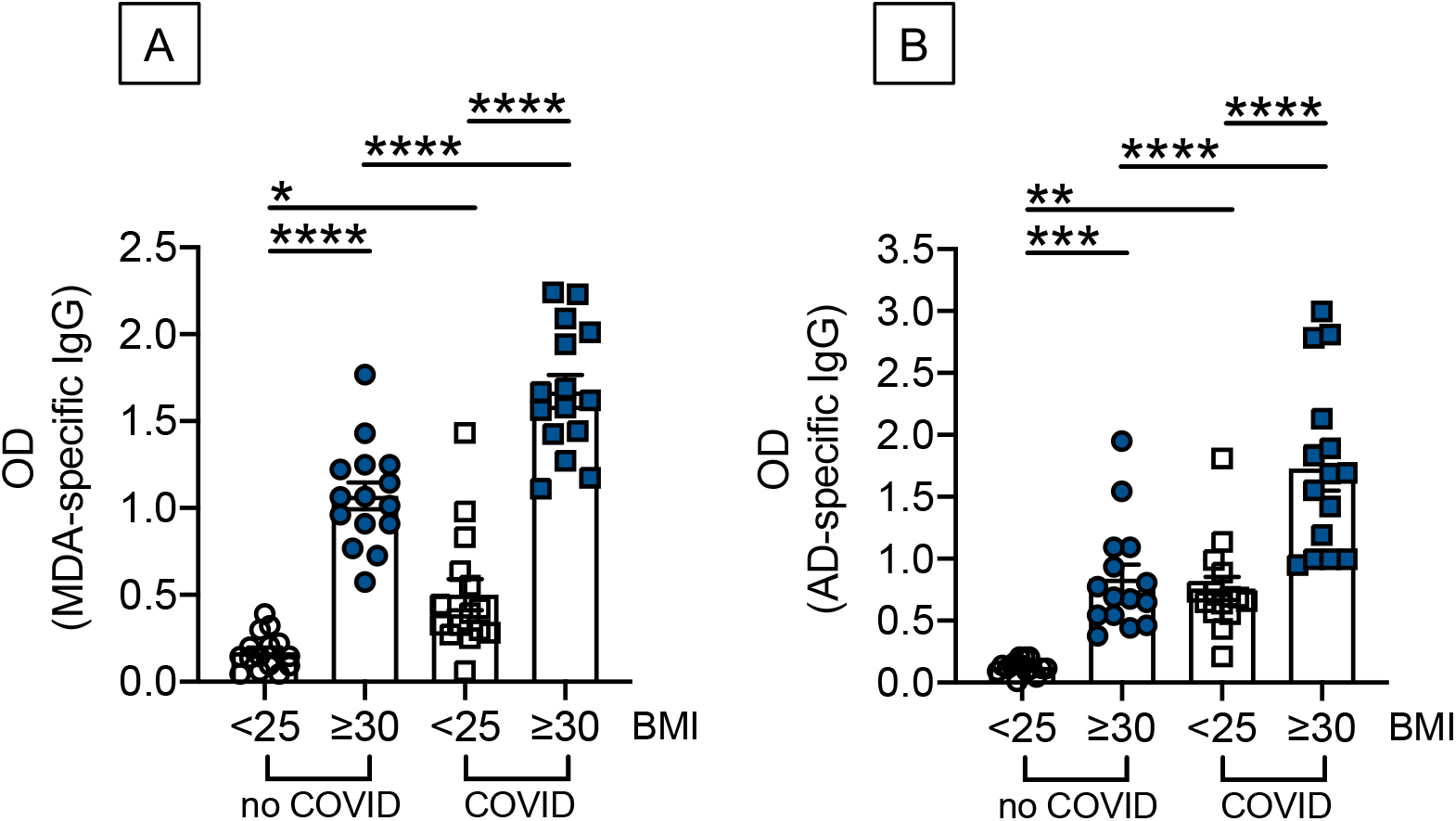
Evaluation of autoimmune IgG antibodies in serum samples of lean and obese COVID-19 patients, as compared to uninfected controls. Anti-MDA (**A**) and anti-AD (**B**) IgG antibodies were measured by ELISA. *p<0.05, **p<0.01, ***p<0.001, ****p<0.0001.

### Association of autoimmune IgG antibodies with CRP

Early reports have shown that autoimmune IgG antibodies in adult COVID-19 patients are correlated with serum levels CRP (36). Therefore, the association of anti-MDA and anti-AD with serum CRP was examined. Results in Fig. 3 show, as expected, higher serum levels of CRP in obese versus lean COVID-19 patients. Results in Fig. 4 show significant associations between CRP serum levels and autoimmune antibodies in our cohort of lean and obese COVID-19 patients.

**Figure 3.**
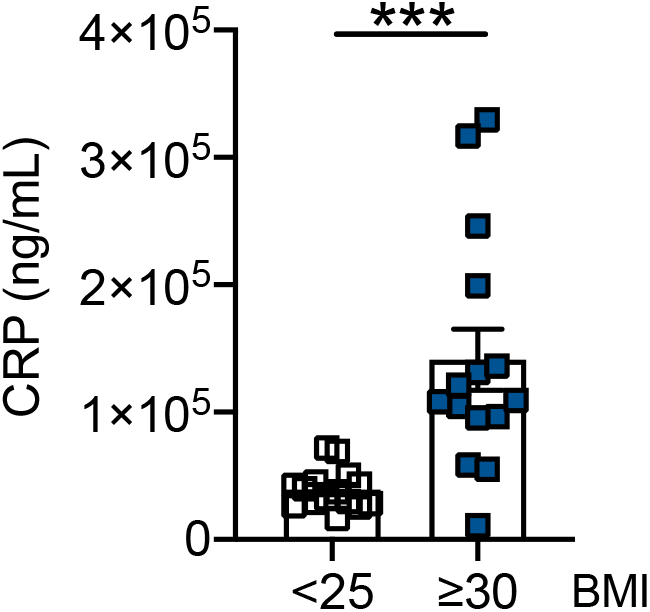
Serum levels of CRP are higher in obese versus lean COVID-19 patients. CRP serum levels were measured by ELISA. ***p<0.001.

**Figure 4.**
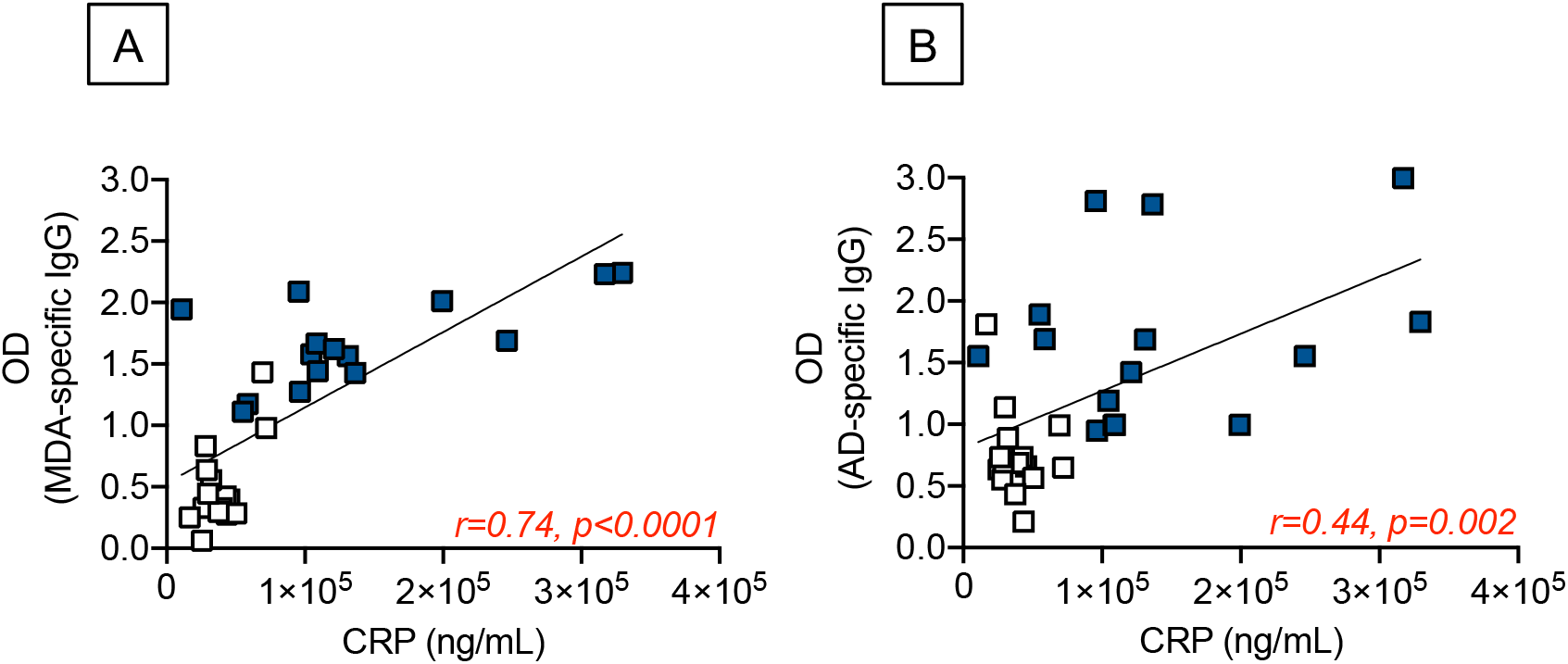
Serum levels of CRP are positively correlated with autoimmune IgG antibodies in lean and obese COVID-19 patients. Correlations of serum CRP with anti-MDA (**A**) and anti-AD (**B**) are shown. Symbol designations as in Fig 3. Pearson’s regression coefficients and p values are indicated at the bottom of each figure.

## Discussion

This study measured serum levels of antibodies with two different autoimmune specificities, MDA and AD, in lean and obese COVID-19 patients compared to lean and obese uninfected controls without previous history of autoimmunity. Obesity has already been shown to be associated with increased oxidative stress and lipid peroxidation (measured by MDA) (42, 43) and increased fat mass (measured by adipocyte-derived proteins released in the AT, AD) (28).

The onset of autoimmunity has been associated with viral infections, and it has been suggested that SARS-CoV-2 could be a triggering factor for the development of a rapid autoimmune, autoinflammatory disease in genetically predisposed individuals as those with high systemic IL-6 (44, 45), similar to what has been found in SARS-CoV, influenza and dengue infections. In SARS-CoV patients, high levels of serum autoantibodies specific for type-2 pneumocytes have been found, and these antibodies were highly cytotoxic (46). In influenza patients, virus-induced autoantibodies against the alveolar and glomerular basement membrane induced the autoimmune disease called Goodpasture’s syndrome (47, 48). In dengue patients, virus-induced autoantibodies specific for endothelial cells, platelets, and coagulatory molecules lead to abnormal activation or dysfunction (49). In all these cases, molecular mimicry between viral and host proteins may explain the cross-reactivity of induced autoantibodies.

In the current pandemic, it has been shown that serum samples of adult COVID-19 patients contain antibodies that target the tissues of infected patients instead of targeting the disease-causing virus. Anti-phospholipid, anti-type-I interferons, anti-nuclear antibodies, and Rheumatoid Factor have been found in a large number of COVID-19 patients and linked to severe disease as they may inactivate critical components of the anti-viral response (50). These findings have suggested that SARS-CoV-2 infection, following dissemination of the virus through the blood, may induce severe tissue damage, cell death, and release of intracellular self-antigens not previously known as disease-causing autoantigens, leading to a self-tolerance breakdown. We cannot exclude the possibility that the virus may also trigger autoimmunity through other mechanisms such as molecular mimicry between viral proteins and self-antigens or reactivation of local viral or bacterial pathogens that have not been removed due to dysfunctional immunity responses in infected patients. We expected the secretion of autoimmune IgG to be increased in obese patients. However, our results have clearly shown that SARS-CoV-2 infection significantly increases serum levels of anti-MDA and anti-AD IgG antibodies more in lean than in obese patients. Serum levels of these antibodies, however, were always higher in obese versus lean COVID-19 patients. These results suggest that the infection has induced de novo autoimmune responses and self-tolerance breakdown following severe tissue damage in lean patients and demonstrates a heightened response in obese patients in which these processes are already occurring.

We also measured serum Spike-specific IgG antibodies in our cohorts of lean and obese COVID-19 patients and uninfected controls. These antibodies were found to be significantly lower in obese as compared to lean COVID-19 patients, confirming our previously published findings that BMI is negatively associated with anti-Spike IgG serum levels (33) and consistent with the knowledge that obesity is associated with inflammaging (2) and metaflammation (51) both of which are negatively associated with a functional immune system (52). Our previously published work has shown that overweight/obesity decrease the serum antibody response to the influenza vaccine in young and elderly individuals (12). As expected, serum Spike-specific IgG antibodies were not found in lean and obese age-gender- and BMI-matched controls.

In conclusion, our results highlight the importance of identifying autoimmune antibodies and biomarkers of self-tolerance breakdown in COVID-19 patients with obesity. Similar autoimmune antibodies may also be secreted following COVID-19 vaccination. However, the reactogenicity of lipid nanoparticle-formulated COVID-19 mRNA vaccines in individuals with obesity, characterized by dysregulation of immune responses, has not been investigated yet. Evaluating the quality of the antibody response of obese COVID-19 patients is crucial to protect this vulnerable population at higher risk of responding poorly to infection with SARS-CoV-2 and vaccination against SARS-CoV-2 lean controls.

## Data Availability

All data are available upon request to the corresponding author.

## Acknowledgments

This study was supported by NIH awards AG32576, AG059719, AG023717, and the University of Miami Department of Pathology & Laboratory Medicine.

## Competing interests

The authors have nothing to disclose.

## Notes

### Competing Interest Statement

The authors have declared no competing interest.

### Author Declarations

The research has been approved and reviewed by the Institutional Review Board (IRB, protocol #20200504, PI Dr. Reidy, and protocols #20070481 and #20160542, PI Dr. Frasca) at the University of Miami, which reviews all human research conducted under the auspices of the University of Miami.

## References

1. Henderson LA, Canna SW, Schulert GS, Volpi S, Lee PY, Kernan KF, et al. On the Alert for Cytokine Storm: Immunopathology in COVID-19. Arthritis Rheumatol. 2020;72(7):1059–63.

2. Franceschi C, Bonafe M, Valensin S, Olivieri F, De Luca M, Ottaviani E, et al. Inflamm-aging. An evolutionary perspective on immunosenescence. Ann N Y Acad Sci. 2000;908:244–54.

3. Mueller AL, McNamara MS, Sinclair DA. Why does COVID-19 disproportionately affect older people? Aging (Albany NY). 2020;12(10):9959–81.

4. Frasca D, Diaz A, Romero M, Garcia D, Blomberg BB. B Cell Immunosenescence. Annu Rev Cell Dev Biol. 2020;36:551–74.

5. Newton AH, Cardani A, Braciale TJ. The host immune response in respiratory virus infection: balancing virus clearance and immunopathology. Semin Immunopathol. 2016;38(4):471–82.

6. Casadevall A, Pirofski LA. The convalescent sera option for containing COVID-19. J Clin Invest. 2020;130(4):1545–8.

7. Duan K, Liu B, Li C, Zhang H, Yu T, Qu J, et al. Effectiveness of convalescent plasma therapy in severe COVID-19 patients. Proc Natl Acad Sci U S A. 2020;117(17):9490–6.

8. Langhi DM, Santis GC, Bordin JO. COVID-19 convalescent plasma transfusion. Hematol Transfus Cell Ther. 2020;42(2):113–5.

9. Falagas ME, Kompoti M. Obesity and infection. Lancet Infect Dis. 2006;6(7):438–46.

10. Karlsson EA, Beck MA. The burden of obesity on infectious disease. Exp Biol Med (Maywood). 2010;235(12):1412–24.

11. O’Shea D, Corrigan M, Dunne MR, Jackson R, Woods C, Gaoatswe G, et al. Changes in human dendritic cell number and function in severe obesity may contribute to increased susceptibility to viral infection. Int J Obes (Lond). 2013;37(11):1510–3.

12. Frasca D, Ferracci F, Diaz A, Romero M, Lechner S, Blomberg BB. Obesity decreases B cell responses in young and elderly individuals. Obesity (Silver Spring). 2016;24(3):615–25.

13. Ovsyannikova IG, White SJ, Larrabee BR, Grill DE, Jacobson RM, Poland GA. Leptin and leptin-related gene polymorphisms, obesity, and influenza A/H1N1 vaccine-induced immune responses in older individuals. Vaccine. 2014;32(7):881–7.

14. Sheridan PA, Paich HA, Handy J, Karlsson EA, Hudgens MG, Sammon AB, et al. Obesity is associated with impaired immune response to influenza vaccination in humans. Int J Obes (Lond). 2012;36(8):1072–7.

15. George MD, Baker JF. The Obesity Epidemic and Consequences for Rheumatoid Arthritis Care. Curr Rheumatol Rep. 2016;18(1):6.

16. Hotamisligil GS. Inflammation and metabolic disorders. Nature. 2006;444(7121):860–7.

17. Johnson AM, Olefsky JM. The origins and drivers of insulin resistance. Cell. 2013;152(4):673–84.

18. Shoelson SE, Lee J, Goldfine AB. Inflammation and insulin resistance. J Clin Invest. 2006;116(7):1793–801.

19. Apovian CM, Gokce N. Obesity and cardiovascular disease. Circulation. 2012;125(9):1178–82.

20. Renehan AG, Tyson M, Egger M, Heller RF, Zwahlen M. Body-mass index and incidence of cancer: a systematic review and meta-analysis of prospective observational studies. Lancet. 2008;371(9612):569–78.

21. Casas R, Sacanella E, Estruch R. The immune protective effect of the Mediterranean diet against chronic low-grade inflammatory diseases. Endocr Metab Immune Disord Drug Targets. 2014;14(4):245–54.

22. Setty AR, Curhan G, Choi HK. Obesity, waist circumference, weight change, and the risk of psoriasis in women: Nurses’ Health Study II. Arch Intern Med. 2007;167(15):1670–5.

23. Naderali EK, Ratcliffe SH, Dale MC. Obesity and Alzheimer’s disease: a link between body weight and cognitive function in old age. Am J Alzheimers Dis Other Demen. 2009;24(6):445–9.

24. Singh-Manoux A, Dugravot A, Shipley M, Brunner EJ, Elbaz A, Sabia S, et al. Obesity trajectories and risk of dementia: 28 years of follow-up in the Whitehall II Study. Alzheimers Dement. 2018;14(2):178–86.

25. Hass DJ, Brensinger CM, Lewis JD, Lichtenstein GR. The impact of increased body mass index on the clinical course of Crohn’s disease. Clin Gastroenterol Hepatol. 2006;4(4):482–8.

26. Ritter A, Kreis NN, Louwen F, Yuan J. Obesity and COVID-19: Molecular Mechanisms Linking Both Pandemics. Int J Mol Sci. 2020;21(16).

27. Lighter J, Phillips M, Hochman S, Sterling S, Johnson D, Francois F, et al. Obesity in Patients Younger Than 60 Years Is a Risk Factor for COVID-19 Hospital Admission. Clin Infect Dis. 2020;71(15):896–7.

28. Frasca D, Diaz A, Romero M, Thaller S, Blomberg BB. Secretion of autoimmune antibodies in the human subcutaneous adipose tissue. PLoS One. 2018;13(5):e0197472.

29. Grant RW, Dixit VD. Adipose tissue as an immunological organ. Obesity (Silver Spring). 2015;23(3):512–8.

30. Franssen FM, O’Donnell DE, Goossens GH, Blaak EE, Schols AM. Obesity and the lung: 5. Obesity and COPD. Thorax. 2008;63(12):1110–7.

31. Murugan AT, Sharma G. Obesity and respiratory diseases. Chron Respir Dis. 2008;5(4):233–42.

32. Kruglikov IL, Scherer PE. The role of adipocytes and adipocyte-like cells in the severity of COVID-19 infections. Obesity (Silver Spring). 2020; https://doi.org/10.1002/oby.22856.

33. Frasca D, Reidy L, Cray C, Diaz A, Romero M, Kahl K, et al. Influence of obesity on serum levels of SARS-CoV-2-specific antibodies in COVID-19 patients. PLoS One. 2021;16(3):e0245424.

34. Frasca D, Diaz A, Romero M, Blomberg BB. Phenotypic and Functional Characterization of Double Negative B Cells in the Blood of Individuals With Obesity. Front Immunol. 2021;12:616650.

35. Frasca D, Diaz A, Romero M, Garcia D, Jayram D, Thaller S, et al. Identification and Characterization of Adipose Tissue-Derived Human Antibodies With “Anti-self” Specificity. Front Immunol. 2020;11:392.

36. Woodruff M, Ramonell R, Lee E-H, Sanz I. Clinically identifiable autoreactivity is common in severe SARS-CoV-2 Infection. medRxiv. 2020.

37. Luo X, Zhou W, Yan X, Guo T, Wang B, Xia H, et al. Prognostic value of C-reactive protein in patients with COVID-19. Clin Infect Dis. 2020.

38. Manson JJ, Crooks C, Naja M, Ledlie A, Goulden B, Liddle T, et al. COVID-19-associated hyperinflammation and escalation of patient care: a retrospective longitudinal cohort study. Lancet Rheumatol. 2020;2(10):e594–e602.

39. Velavan TP, Meyer CG. Mild versus severe COVID-19: Laboratory markers. Int J Infect Dis. 2020;95:304–7.

40. Wang L. C-reactive protein levels in the early stage of COVID-19. Med Mal Infect. 2020;50(4):332–4.

41. Bradford MM. A rapid and sensitive method for the quantitation of microgram quantities of protein utilizing the principle of protein-dye binding. Anal Biochem. 1976;72:248–54.

42. Sankhla M, Sharma TK, Mathur K, Rathor JS, Butolia V, Gadhok AK, et al. Relationship of oxidative stress with obesity and its role in obesity induced metabolic syndrome. Clin Lab. 2012;58(5-6):385–92.

43. Yesilbursa D, Serdar Z, Serdar A, Sarac M, Coskun S, Jale C. Lipid peroxides in obese patients and effects of weight loss with orlistat on lipid peroxides levels. Int J Obes (Lond). 2005;29(1):142–5.

44. Caso F, Costa L, Ruscitti P, Navarini L, Del Puente A, Giacomelli R, et al. Could Sars-coronavirus-2 trigger autoimmune and/or autoinflammatory mechanisms in genetically predisposed subjects? Autoimmun Rev. 2020;19(5):102524.

45. Gagiannis D, Steinestel J, Hackenbroch C, Schreiner B, Hannemann M, Bloch W, et al. Clinical, Serological, and Histopathological Similarities Between Severe COVID-19 and Acute Exacerbation of Connective Tissue Disease-Associated Interstitial Lung Disease (CTD-ILD). Front Immunol. 2020;11:587517.

46. Lin YS, Lin CF, Fang YT, Kuo YM, Liao PC, Yeh TM, et al. Antibody to severe acute respiratory syndrome (SARS)-associated coronavirus spike protein domain 2 cross-reacts with lung epithelial cells and causes cytotoxicity. Clin Exp Immunol. 2005;141(3):500–8.

47. Gunnarsson A, Hellmark T, Wieslander J. Molecular properties of the Goodpasture epitope. J Biol Chem. 2000;275(40):30844–8.

48. Reynolds J, Moss J, Duda MA, Smith J, Karkar AM, Macherla V, et al. The evolution of crescentic nephritis and alveolar haemorrhage following induction of autoimmunity to glomerular basement membrane in an experimental model of Goodpasture’s disease. J Pathol. 2003;200(1):118–29.

49. Wan SW, Lin CF, Yeh TM, Liu CC, Liu HS, Wang S, et al. Autoimmunity in dengue pathogenesis. J Formos Med Assoc. 2013;112(1):3–11.

50. Woodruff MC, Ramonell RP, Lee FE, Sanz I. Clinically identifiable autoreactivity is common in severe SARS-CoV-2 Infection. https://www.medrxivorg/content/101101/2020102120216192v2. 2020.

51. Hotamisligil GS. Inflammation, metaflammation and immunometabolic disorders. Nature. 2017;542(7640):177–85.

52. Mayer-Pickel K. Obesity and Antiphospholipid Syndrome: A Particular Challenge in Pregnancy. Obesity Research. 2015;2(2):46–56.

